# Polygenic proxies of age-related plasma protein levels reveal TIMP2 role in cognitive performance

**DOI:** 10.1101/2024.07.23.24310854

**Authors:** Federica Anastasi, Patricia Genius, Blanca Rodriguez-Fernandez, Chengran Yang, Priyanka Gorijala, Jigyasha Timsina, Felipe Hernández-Villamizar, Luigi Lorenzini, Marta del Campo, Gonzalo Sánchez-Benavides, Carolina Minguillon, Arcadi Navarro, Carlos Cruchaga, Marc Suárez-Calvet, Natalia Vilor-Tejedor

## Abstract

Several studies have identified blood proteins that influence brain aging performance in mice, yet translating these findings to humans remains challenging. Here we found that higher predicted plasma levels of Tissue Inhibitor of Metalloproteinases 2 (TIMP2) were significantly associated with improved global cognition and memory performance in humans. We first identified 12 proteins with aging or rejuvenating effects on murine brains through a systematic review. Using protein quantitative trait loci data for these proteins, we computed polygenic scores as proxies for plasma protein levels and validated their prediction accuracy in two independent cohorts. Association models between genetic proxies and cognitive performance highlighted the significance of TIMP2, also when the models were stratified by sex, *APOE*-ε4, and Aβ42 status. This finding aligns with TIMP2’s brain-rejuvenating role in murine models, suggesting it as a promising therapeutic target for brain aging and age-related brain diseases in humans.

## 1. INTRODUCTION

Aging is the main risk factor for Alzheimer’ disease (AD) and most neurodegenerative diseases. Despite this well-established link, the underlying mechanisms connecting aging with AD remain unknown and, up to now, potential therapies for AD by targeting aging have been poorly explored^1–4^. A promising avenue of research has emerged from studies on murine models revealing the presence of circulation blood factors that either accelerate or slow the aging of multiple organs including the brain^5–18^. These blood factors have predominantly been identified through parabiosis experiments, in which a young and an old mouse undergo a surgical technique that joins the circulatory system of these two living animals^5,11,13,18,19^. The effect on aging of some of these blood factors identified by parabiosis have been confirmed by their systemic injection. Several circulating blood factors have been described in mice to have an aging effect on the brain or, conversely, slowing or reversing the cellular hallmarks of aging, even improving in such instances the performance in cognitive and behavioral tests. These latter circulating blood factors include proteins such as the Tissue Inhibitor of Metalloproteinases 2 (TIMP2) that has been described as “rejuvenating”^13^. From these studies, it could be speculated that targeting these age-related circulating factors (by promoting or counteracting their effects) may serve as a therapeutic strategy to ameliorate age-related cognitive deficits, as those occurring in AD. Whether these circulating proteins have a similar effect in humans as observed in animal models is unknown, and despite these promising results, the translatability of these findings to humans has yet to be clearly elucidated. Observational studies in humans have explored the association between the levels of circulating blood factors previously described in mice with cognition or neurodegeneration markers. For example, a study found that the simultaneous increase of the pro-aging blood factors CCL2 and CCL11 in AD dementia patients predicted worse verbal and visual memory scores^20^. Another study found that CCL2 in blood is associated with faster cognitive decline in early AD dementia^21^. It is crucial to better investigate whether the age-related circulating blood factors identified in mice similarly affect the human brain, as these could be therapeutically targeted to mitigate age-related neurological conditions.

The present study introduces an innovative approach by integrating proteomic quantitative trait loci (pQTL) into protein-based polygenic risk scores (protPRS) to address these limitations, facilitating the translation of findings from animal models to humans. By leveraging genetically predicted plasma protein levels implicated in aging and rejuvenation processes, this study aimed to assess the effect of these blood factors on cognitive performance in humans at risk of AD, potentially offering a more immediate reflection of the proteome’s impact on neurodegeneration. We first performed a systematic review to identify those circulating blood factors that have been described in murine models to have an aging or rejuvenating effect on the brain. Next, we computed protPRS for these identified circulating blood factors, incorporating genetic variants with established associations to protein expression levels. Finally, following the validation of these protPRS against actual protein level measurements in two independent samples, we explored the relationship between the protPRS, serving as a proxy for actual protein levels, and cognitive performance metrics in cognitively unimpaired (CU) individuals at higher risk for AD dementia (Figure 1).

**Figure 1.**
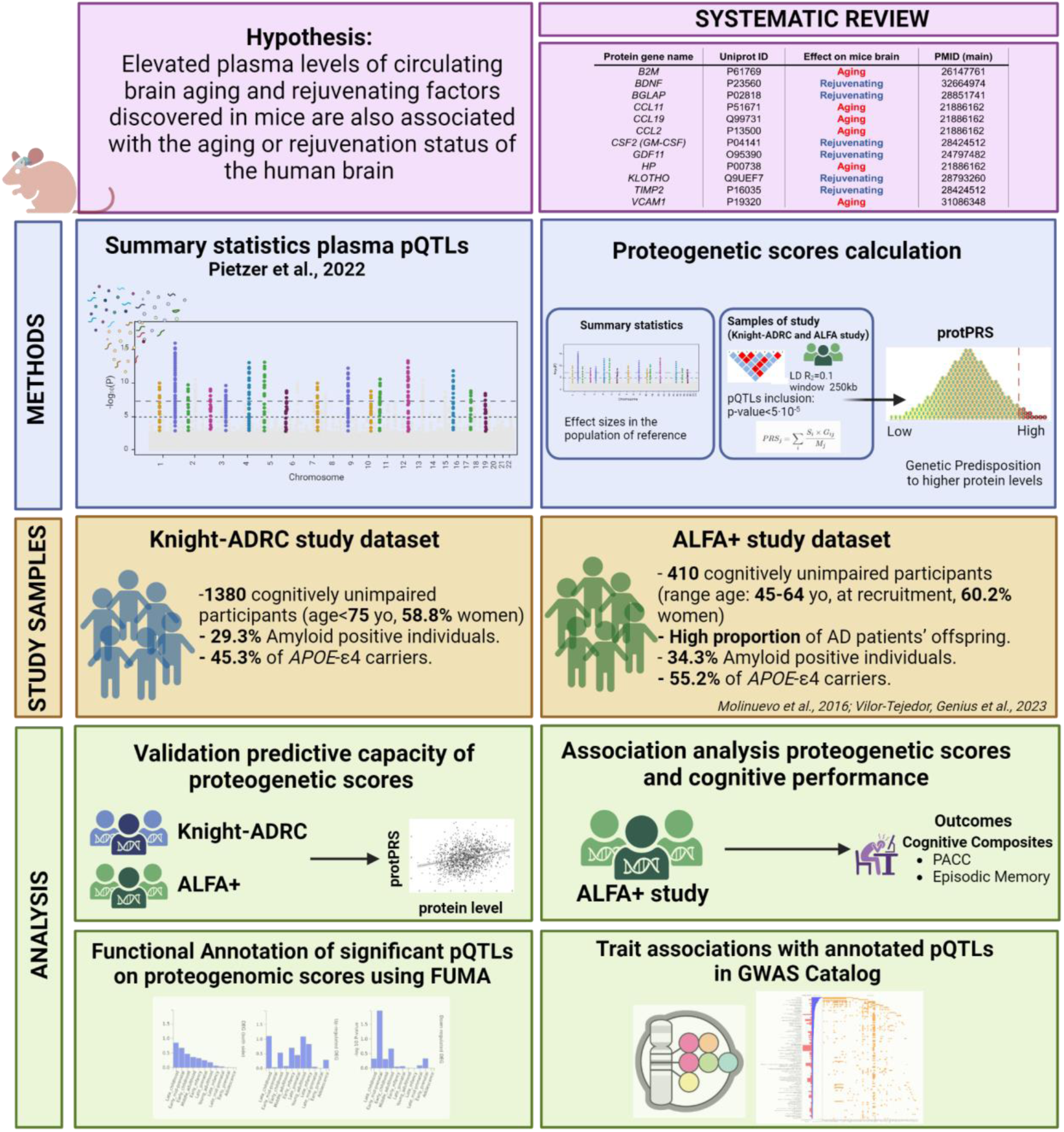
Workflow of the study.

## 2. RESULTS

### 2.1 Identification of circulating blood proteins with an aging or rejuvenating effect: a systematic review

Following a systematic review adhering to PRISMA guidelines^22^, we identified 21 publications that collectively encompassed 17 proteins linked to aging or rejuvenating effects in mice (Supplementary Tables 1 and 2). Inclusion, exclusion and search criteria are described in the Methods section. A diagram of the full systematic review workflow is available in Supplementary Figure 1. In this study, we use the term “rejuvenation” to describe the process of reversing cellular hallmarks of aging and restoring cognitive function to a more youthful state^23^. Since the primary focus of our research is to delve into the potential relationship between these identified proteins and their role in human brain aging and cognitive impairment, we selectively extracted from the systematic review those proteins with direct implications on brain function. The final list of the 12 brain-relevant proteins considered for this study is detailed in Table 1.

**Table 1.**
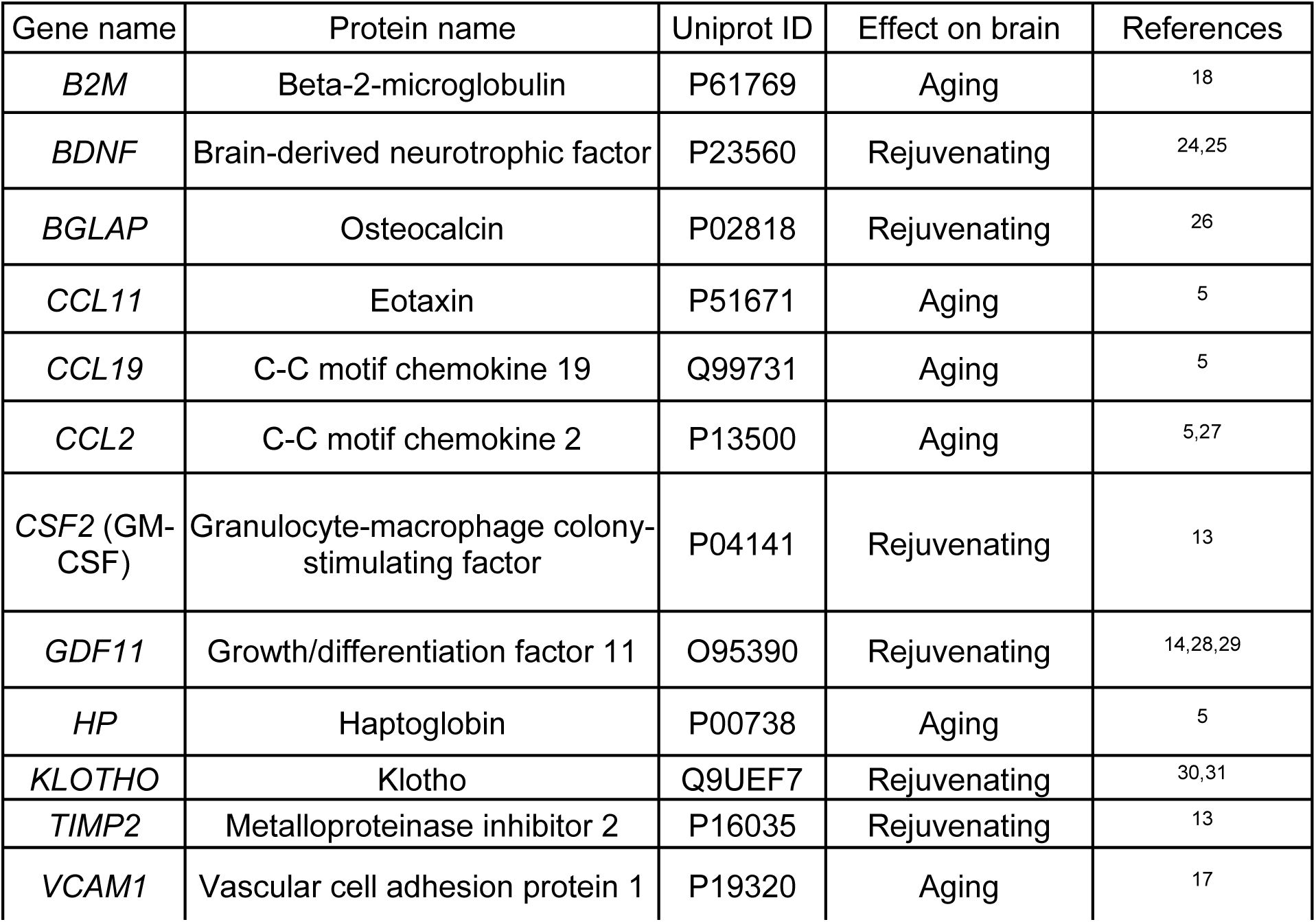
Circulating blood proteins influencing mice brain aging identified through systematic review. For each identified protein, the table provides identifiers including the *Gene name, protein name and UniProt ID.* The table additionally includes the reported effect on the mouse brain (aging/rejuvenating) and the references to the respective papers included in the systematic review.

### 2.2 Polygenic protein scores as proxies of plasma protein levels

We computed a total of 13 protPRSs. Two proteins (BDNF and HP) had available summary statistics data for two different measurements of the same protein. For CCL11 no summary statistics from the Fenland study were available (Supplementary Table 3). The protPRSs were significantly associated with the actual plasma protein levels for 10 and 7 out of the 13 protPRSs in the Knight-ADRC cohort and ALFA+ cohort, respectively (Table 2, Supplementary Figure 2-3). The protPRSs of HP (both scores), KLOTHO, B2M, VCAM1 and TIMP2 consistently predicted their respective protein levels in both cohorts. Conversely, the protPRSs of CCL2, and CSF2 did not exhibit a significant association with their respective protein levels in none of the cohorts. Although the GDF11 protPRS significantly predicted GDF11 plasma protein levels in the ALFA+ cohort, it was not considered validated since it was negatively associated (ꞵ<0) with the GDF11 plasma protein levels in the Knight-ADRC cohort. Of note, HP protPRS elucidates a substantial portion of the variability in HP protein levels, with protPRSs-R^2^ values of approximately 30% in the Knight-ADRC cohort, and 12% in the ALFA+ cohort (Table 2). Due to the low statistical power in the ALFA+ cohort after multiple testing correction, we considered as valid protPRSs those that, at least, significantly predicted the actual plasma protein levels in the Knight-ADRC cohort.

**Table 2.** protPRSs association with plasma protein levels in the Knight-ADRC and ALFA+ cohorts. Outcomes of linear association models between each of the protPRS and its corresponding plasma protein. Models were adjusted for age and sex. For each protPRS, the standardized effect, R-squared (genetic components) and FDR-corrected p-values of the association models are provided.

|  | Knight-ADRC |  |  | ALFA+ |  |  |
| --- | --- | --- | --- | --- | --- | --- |
| protPRS-protein pair | St. beta | protPRS – $R^2$ (%) | FDR adj. P-value | St. beta | protPRS – $R^2$ (%) | FDR adj. P-value |
| B2M | 0.289 | 8.7% | **2.41E-26** | 0.233 | 5.4% | **3.21E-05** |
| BDNF (1) | 0.091 | 1% | **0.001** | 0.046 | 0.2% | 0.556 |
| BDNF (2) | 0.070 | 0.5% | **0.013** | -0.015 | 0.02% | 0.842 |
| BGLAP | 0.115 | 1.4% | **3.13E-05** | -0.002 | 0.00% | 0.973 |
| CCL19 | 0.142 | 1.9% | **5.56E-07** | 0.110 | 1.2% | 0.085 |
| CCL2 | 0.046 | 0.2% | 0.080 | -0.030 | 0.1% | 0.689 |
| GDF11 | -0.028 | 0.1% | 0.340 | 0.143 | 2.1% | 0.017 |
| CSF2 | 0.038 | 0.2% | 0.185 | -0.031 | 0.1% | 0.689 |
| HP (1) | 0.583 | 34.2% | **1.09E-119** | 0.383 | 11.8% | **1.20E-12** |
| HP (2) | 0.555 | 30.3% | **4.90E-103** | 0.379 | 12.2% | **1.20E-12** |
| KLOTHO | 0.263 | 7% | **1.93E-22** | 0.252 | 5.7% | **1.93E-05** |
| TIMP2 | 0.122 | 1.6% | **3.50E-06** | 0.140 | 1.8% | **0.017** |
| VCAM1 | 0.161 | 2.9% | **1.85E-09** | 0.130 | 1.6% | **0.024** |

### 2.3. TIMP2 protPRS is associated with cognitive performance in human individuals at risk of Alzheimer’s disease

To investigate whether the circulating blood proteins linked to brain aging in murine models have a similar effect in humans, we studied the association between validated protPRS and cognitive performance in CU individuals of the ALFA+ cohort at risk of AD dementia.

Results revealed that genetic predisposition to elevated plasma levels of TIMP2, a protein that was described as having a rejuvenating effect on the mice brain^13^, was associated with better cognitive performance as measured by the Preclinical Alzheimer Cognitive Composite (PACC), and episodic memory (EM) composite score. The association of TIMP2 protPRSs with better PACC was also significant in models stratified by sex, Aβ status (as defined by CSF Aβ42/40) and *APOE-*ε4 carriership. After stratifying for Aβ status, significance did not survive FDR p-value correction for multiple comparisons, although the association was significant at nominal level and group-correction, respectively (Figure 2, Supplementary Table 4-5).

**Figure 2.**
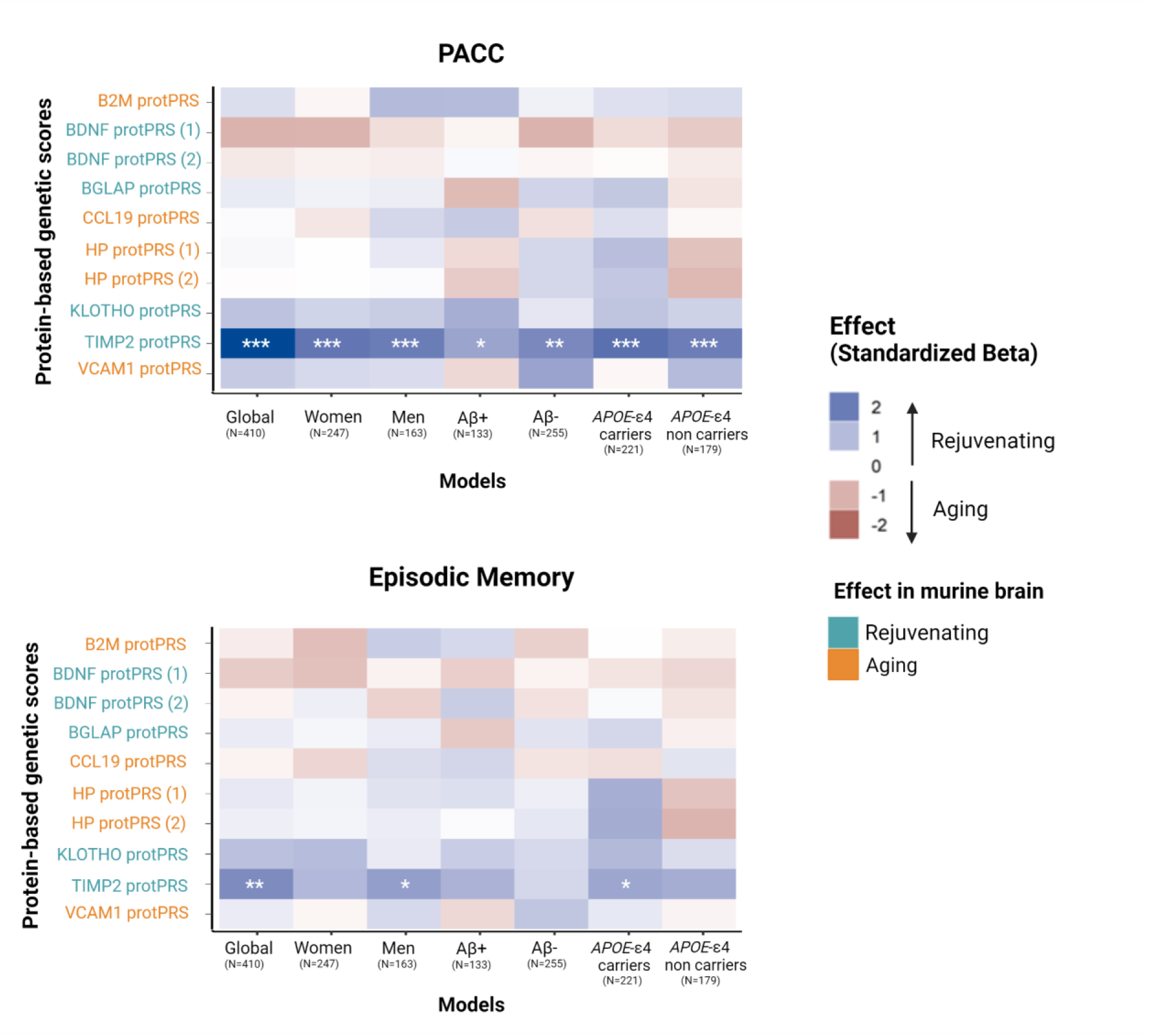
Results of the association analysis of the computed protein-based genetic scores (protPRSs) with the cognitive composites PACC and EM. Each box represents an association model and colors represent the direction of the association effect. Blue refers to a rejuvenating effect and red to an aging effect in mice. Significance is reported at nominal level, group-correction (two experimental aging groups) and FDR-multiple correction (*p<0.05, **p<0.025, ***FDR-corrected p-value <0.05 respectively). protPRS label colors represent the reported effect in murine studies, where light blue indicates a rejuvenating effect and orange indicates an aging effect. BDNF and HP have summary statistics for two different aptamers. Legend: Aβ+ (amyloid positive individuals defined by CSF Aβ42/40≤0.071); Aβ-(amyloid positive individuals defined by CSF Aβ42/40>0.071), *APOE*-ε4 carriers (carriers of at least one copy of *APOE*-ε4).

We further performed a cross-platform robustness check, measuring TIMP2 in ALFA+ with an ELISA immunoassay. TIMP2 levels measured by ELISA immunoassay correlates with those measured from the SomaScan technology, confirming the SomaScan aptamer’s effective recognition of the TIMP2 protein (Supplementary Figure 4). Moreover, TIMP2 protPRS was correlated at a nominal level with TIMP2 levels measured by ELISA (Supplementary Figure 5). These findings validate that the TIMP2 protPRS computed in this study is indeed a robust proxy for the plasma TIMP2 protein. Nevertheless, non-significant results were found between cognitive function (PACC and EM) and the actual TIMP2 levels (as measured by ELISA or the SomaScan platform; see Supplementary Table 6).

### 2.4. Functional mapping, annotation and gene set enrichment analysis of variants included in TIMP2 protPRS

The TIMP2 protPRS, derived from the ALFA+ cohort, included 118 pQTLs significantly associated (p-value<5×10^-5^) with TIMP2 protein levels as identified in the Pietzner et al. study (Supplementary Table 7). Functional annotation analysis revealed (after Bonferroni correction over 11 categories) that TIMP2 protPRS was enriched for intronic (47.7% for pQTLs variants *vs*. 36.4% in a reference panel), exonic (1.4% *vs*. 1%), and 3’-UTR (1.4% *vs*. 0.9%) variants, while intergenic (40.3% *vs*. 46.6%), and intronic non-coding RNA (5.5% *vs*. 11.5%) were significantly less common (Supplementary Figure 6).

In addition, we found that regions within genes on chromosome 3 may be important for regulating the expression of the TIMP2 protein, including the most significant variants annotated as TLR9, SMIM4, STAB1 and RTF1 (Supplementary Figure 7A). Specifically, the variant *rs445676* in the Toll-like receptor 9 (*TLR9*) gene, showed the highest effect, and the variant *rs76685449* in the Small Integral Membrane Protein 4 (*SMIM4*) gene near the *TLR9* gene, was the most statistically significant and had the highest negative effect.

Enrichment analyses showed a significant involvement of annotated variants in several lipid-related gene ontology terms. The most significant associations were found with low-density lipoprotein particle binding. Other relevant functions included protein-lipid complex binding, glycosaminoglycan binding, and heparin binding, suggesting a likely functional role of TIMP2 in lipid metabolism and interaction pathways (Supplementary Figure 7B). Finally, by integrating genetic data with expression across 54 tissues, we found that genes associated with TIMP2 pQTLs exhibited tissue-dependent regulation, which has significant implications for several brain regions (Supplementary Figure 8).

## 3. DISCUSSION

In this study, we showed that the genetically predicted levels of TIMP2 in plasma are associated with better cognition in CU individuals at risk of AD dementia. These results provide a link between the discovery of TIMP2 in murine models as a blood factor with a rejuvenating effect on the mouse brain, and the confirmation in humans of the association between its predicted levels and higher cognitive performance. Translating the discovery of age-related blood factors in humans is challenging^32^. Observational studies have attempted to bridge this gap by measuring the levels of selected circulating blood factors identified in mice and assessing their association with cognition in humans^33^. CCL2 and CCL11 in blood were previously associated with a poorest cognition^21,34,35^ and, more recently, machine learning models were used to define organ-specific blood plasma protein signatures that were associated with accelerated aging, including accelerated brain aging and AD^36^. However, assessing single blood measurements may not accurately reflect lifelong exposure to these factors, further limiting the translatability of the results. Additionally, acquisition of longitudinal information, crucial for an in-depth analysis, involves substantial expenses, both in economic terms and in computational resources.

Here, we propose an alternative and innovative cost-effective approach to circumvent these challenges. We provided evidence for the utility of protPRSs, derived from plasma pQTLs data, in predicting plasma protein levels and exploring their association with cognitive function, particularly in the context of aging. Our findings underscored the significant potential of protPRSs, integrating pQTLs data, for elucidating the complex interplay between genetic predispositions, plasma protein levels, and cognitive impairment within the aging human population. The main advantage of using protPRSs to predict protein levels is the sample size of the currently available pQTL analyses, which thus provides strong statistical evidence of genetic variants associated with the levels of these proteins. Moreover, protPRSs capture the aggregate variance associated with proteins, offering substantial potential to identify molecular pathways that modulate protein levels.

By validating these protPRSs using genotypic and proteomics data from the Knight-ADRC and ALFA+ cohorts, we have shown that a significant proportion of the protPRS are strongly associated with their corresponding protein levels in plasma. It is important to note that more protPRSs have been validated in the Knight-ADRC cohort, likely due to higher statistical power. Notably, although the pQTL data were sourced from the Fenland study, which comprises individuals of European descent, the validation of protPRSs as proxies for protein levels in the Knight-ADRC cohort, predominantly composed of non-Hispanic white individuals from North America, further underscore their robustness and wide applicability. This validation is crucial, as it supports the hypothesis that genetic variants can serve as reliable predictors of protein expression.

Further analyses in an independent cohort (ALFA+), aimed at assessing the translatability of the findings in murine models to humans by testing the associations between the predicted levels of these proteins in plasma and cognitive performance, in individuals at risk of AD dementia. The main results of this study highlighted the association of TIMP2 protPRS with cognitive performance in CU individuals at risk of AD, specifically in global cognition and memory composites. Importantly, the associations with PACC remained significant when stratified by amyloid status or *APOE-*ε4 genotype, indicating that the effects of the TIMP2 protPRS on cognition is not specific to amyloid or *APOE*-related pathways. Upon discovering the significant association of TIMP2 protPRSs with cognition, we extended our investigation to include the validation of TIMP2 protein levels association with cognition in the same ALFA+ plasma sample. Despite the significant associations, our analysis revealed that the observed link between TIMP2 and cognition did not persist when considering the actual protein measurements. Possible explanation is that genetic variants are stable during lifetime, thus protPRSs may better describe the cumulative life-time effect of a predisposition to high or low plasma levels of TIMP2, regardless of presence of Aꞵ pathology or neurodegeneration. Conversely, protPRSs may not fully capture the complexity of the underlying biology and may miss important environmental or epigenetic factors that contribute to plasma protein level. TIMP2 rejuvenating effect on cognition was firstly described by Castellano et al.^13^: the authors found TIMP2 enriched in human cord plasma, young mouse plasma, and young mouse hippocampus. Systemic administration of TIMP2 increased synaptic plasticity and hippocampal-dependent cognition in aged mice. More recently the same group highlighted TIMP2 as a fine regulator of synaptic plasticity and hippocampus-dependent memory through its interactions with the extracellular matrix^37^. TIMP2 belongs to a small family of tissue metalloproteinase inhibitors that includes TIMP1, TIMP3, and TIMP4, all of which target and regulate matrix metalloproteinases (MMPs) activity. While primarily recognized for its inhibitory activity on proteases like matrix metalloproteinases (MMPs), a study by Britton et al. brought attention to TIMP2 non-canonical role. They underscored that TIMP2’s beneficial effects on cognition and neuronal function in mice were not directly mediated through MMP inhibition^38^. Functional analyses in our study suggested that regions within genes at chromosome 3 might be key in regulating TIMP2 plasma protein levels. In particular, the variant *rs445676*, which exhibited the highest positive effect, was found within the *TLR9* gene. *TLR9* belongs to the Toll-like receptor family, which plays a fundamental role in the activation of the innate and adaptive immunity^39^. Of note, TLR9 is highly expressed when microglia are stimulated with Amyloid-β^40^, and recent evidence reported its role in memory function and neurodegeneration^41,42^. Enrichment analyses also underscored a likely functional role of TIMP2 in lipid metabolism and interaction pathways, further confirmed by the presence of the *APOE* genes among the TIMP2 protPRS annotated variants, suggesting broader implications for aging and aging-related disorders. Moreover, our study showed that the expression of annotated genes responsible for regulating TIMP2 levels in the blood exhibits tissue-dependent up-regulations with significant implications for several brain regions. This finding suggests a potential modulation of TIMP2 activity, which could have profound implications for the maintenance of neurological health and the progression of age-related neurodegenerative diseases.

The strengths of our study lie in the use of protPRSs, which offers complementary benefits to quantifying protein abundance. Firstly, genetic predictions allow us to infer an individual’s stable, lifelong propensity towards certain protein expression levels, independent of transient environmental factors or acute physiological conditions. Secondly, incorporating genetic predictions facilitates the study of a wider array of protein levels that may be challenging to measure directly due to assay limitations or the dynamic nature of protein expression, which can vary significantly throughout the day or across different physiological states. Lastly, the integration of actual and genetically predicted protein levels provides a comprehensive approach to understanding the multifaceted nature of protein involvement in cognitive aging. It allows researchers to assess the consistency and robustness of associations across different levels of biological inference, from genetic predisposition to actual protein expression, thereby enriching our understanding of the biological pathways involved. A potential limitation of our study is its cross-sectional design, which constrains the investigation of the association of TIMP2 protPRSs with longitudinal cognitive decline rates. Future research should consider longitudinal study designs to explore the relationship between TIMP2 protPRSs and cognitive decline rates over extended follow-up periods.

In conclusion, we show that the genetically predicted levels of TIMP2 in plasma are associated with better cognition in humans. We also demonstrate that the application of protPRS as a proxy of protein levels can address translational challenges commonly encountered in animal studies. protPRSs show substantial promise for improving early detection and personalized treatment of cognitive decline. By identifying individuals at higher risk due to their genetic predispositions to altered TIMP2 protein levels, early and targeted interventions can be initiated, potentially altering the disease trajectory. These interventions can be precisely tailored to individual molecular profiles, optimizing therapeutic outcomes and possibly delaying or even preventing the onset of cognitive impairment. As research advances, the integration of protPRSs into clinical practice promises to transform the management and treatment of age-related cognitive disorders, leading the way toward a future of personalized medicine.

## 4. METHODS

### 4.1. Systematic Review

We conducted a comprehensive systematic literature review of publications indexed in PubMed (Medline) and Google Scholar until 2023, to identify circulating proteins with demonstrated aging or rejuvenating effects in mice. Our research adhered to a structured review process aligned with the Preferred Reporting Items for Systematic Reviews and Meta-Analyses (PRISMA) guidelines^22^. The central research question was formulated as follows: “*Which circulating blood proteins have been experimentally shown to manifest either aging or rejuvenating effects on the mouse brain?*”

To address this question, we established specific inclusion and exclusion criteria:

#### Inclusion criteria

1. Studies that used experimental methodologies such as parabiosis, blood administration from young or aged mice (or *vice versa*), or *in vivo* peripheral administration of proteins. Peripheral administration methods encompassed intravenous and subcutaneous routes.
2. Studies assessing aging or rejuvenating effects through assessments of cellular hallmarks of aging. Rejuvenation, in this context, was defined as the process of reversing aging to a more youthful state in terms of organ performance or overall lifespan extension.
3. Peer-reviewed article, written in English, original research studies-only and full-text available.

#### Exclusion criteria

1. Studies involving genetic interventions to either increase or decrease protein levels.
2. Studies conducted on disease models (only effects on healthy animals were considered).
3. Studies that utilized antibody administration to block or reduce the effects of the protein of interest.
4. Studies employing analogs of the protein of interest to simulate its effects.
5. Studies involving the administration of proteins that are not physiologically secreted.

Our literature search was systematically performed using Google Scholar and PubMed, employing the following combinations of keywords: “*lifespan AND (aging OR ageing OR aged) AND mice*”, “*lifespan AND rejuvenating AND mice*”, “*(parabiosis OR parabiose) AND rejuvenating*”, “*(parabiosis OR parabiose) AND (aging OR ageing OR aged)*”, “*(plasma OR serum OR blood) AND rejuvenating AND factor*”, “*(plasma OR serum OR blood) AND (aging OR ageing OR aged) AND factor*”.

Initially, we screened the title and abstract to discard irrelevant studies. We also checked the reference lists of included articles to identify additional relevant publications pertinent to our research question. Using Mendeley Cite as reference manager, organized the pre-selected manuscripts in a dedicated folder, and duplicates were removed. Finally, for each paper we screened the full-text to compile a definitive list of manuscripts that adhered to the aforementioned inclusion and exclusion criteria (Supplementary Figure 1).

For the present study, our primary focus is to investigate the potential associations between proteins known to have an aging or rejuvenating effect on the mouse brain and the process of brain aging and cognitive impairment. Therefore, we have selectively extracted from the review only those proteins that exhibit an effect on the brain.

### 4.2. The Knight-ADRC cohort, genotyping profiling and proteomic assessment

#### Sample description

To validate the utility of the protPRSs as genetic proxies for plasma protein levels, we used proteomics and genetics data from the Charles F. and Joanne Knight Alzheimer Disease Research Center (Knight ADRC) cohort. The study was approved by the Institutional Review Board at Washington University School of Medicine in St. Louis. For the analysis, we selected a subset of 1,380 CU participants, who were less than 70 years old. This specific participant subset was selected to enhance congruence with the demographic characteristics of the Fenland study cohort, from which the summary statistics of the plasma pQTLs was extracted. Comprehensive sociodemographic and clinical information for the entire sample is presented in Table 3.

**Table 3.**
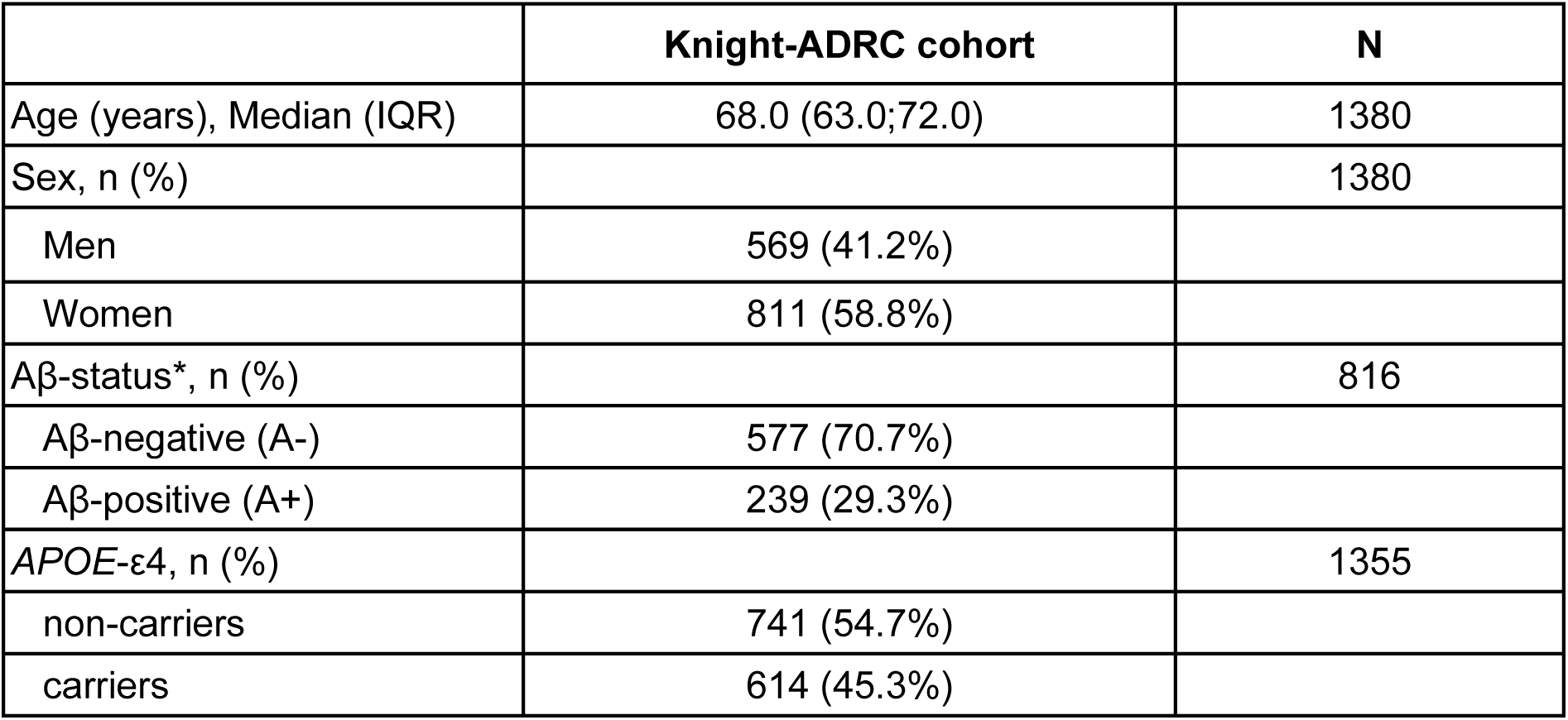
Demographic characteristics of the Knight-ADRC study cohort. Data are reported as percentage (%) [sex, Aβ-status, *APOE*-ε4 carriership] and median (M) and interquartile range (IQR) [age]. Legend: *Aβ positivity (A+) is defined by CSF Aβ42 < 635 pg/ml^43^ or high amyloid probability score (APS)^44^. Abbreviations: Aβ, β-amyloid; *APOE*, apolipoprotein E.

#### Genotyping Profiling

Genotyping was performed using multiple genotyping arrays (CoreEx, GSA_v1, GSA_v2, GSA_v3, NeuroX2, OmniEx, Quad660). For each genotyping array, pre-imputation, imputation and post-imputation were performed separately. After post-imputation, all arrays were merged into one dataset. At the pre-imputation stage, quality control was performed on both variants and sample using PLINK v1.9. Genotyped variants were kept agreeing the following quality control criteria: (1) genotyping successful rate ≥ 98% per variant or per individual; (2) MAF ≥ 0.01; and (3) Hardy–Weinberg equilibrium (HWE) (P ≥ 1×10^−6^). Sample-level quality control included verification of sex codes (filtering out sex mismatches) and the identification of sample ID duplications.

For phasing and imputation, TOPMed server with genome build GRCh38 (imputation panel version R2), imputation quality of R^2^ ≥ 0.3, and Eagle v2.4 phasing was used^45^. In the post-imputation phase, both genotyped and imputed variants were retained based on two criteria: (1) a genotyping missing rate of ≤ 90% per variant; (2) a MAF of ≥ 0.0005.

#### Plasma sample collection and proteomics assessment

The plasma sample proteomics assessment in the Knight-ADRC cohort was extensively discussed in Timsina et al.^46^ In brief, 3,132 participants and 6,907 aptamers passed proteomics QC. 7,584 aptamers were measured before proteomics QC using the SOMAscan 7k platform. Plasma proteomics data from all genetic ancestries were QCed with seven steps: Step 1) Limit of detection, scale factor difference, and coefficient of variation; Step 2) IQR-based outlier expression level detections; Step 3) Remove Analytes and Samples with <65% call rate; Step 4) Re-calculate call rate for analytes and remove analytes with call rate <85%; Step 5) Re-calculate missing rate for subjects and remove subjects with < 85% call rate cut-off; Step 6) Back transformation into raw values; Step 7) Removal of Non-Human and analytes without protein targets and Final matrix.

### 4.3. The ALFA+ study, genotyping profiling, protein quantification and cognitive assessment

#### Sample description

We computed the protPRS in a total of 410 participants of the ALFA+ cohort, a nested longitudinal study of the ALFA parent cohort (Alzheimer and Families) with genotype and cognitive assessment data available^47^. The ALFA parent cohort was established as a research platform to understand the early pathophysiological alterations in preclinical AD and is composed of CU individuals (between 45 and 75 years at baseline), enriched for family history of AD and genetic risk factors for AD^48^. In the present study in the ALFA+ cohort, all individuals were CU and within an age range of 45 to 65 years. 60% of the cohort were women, 34% were positive for CSF β-amyloid (Aβ) status, and 55% were identified as carriers of the *APOE-ε4* allele. CSF β-amyloid (Aβ) status was defined by the CSF Aβ42/40 ratio, and participants were classified as CSF Aβ-positive (A+) if CSF Aβ42/40 ≤ 0.071^49^. Comprehensive sociodemographic and clinical information for the entire sample is presented in Table 4.

**Table 4.** Demographic characteristics of the ALFA+ study cohort. Data are reported as percentage (%) [sex, Aβ-status, *APOE*-ε4 carriership] and median (M) and interquartile range (IQR) [age at lumbar puncture, years of education, PACC and EM]. *Aβ positivity (A+) is defined by CSF Aβ42/40 ≤ 0.071. Abbreviations: Aβ, β-amyloid; *APOE*, apolipoprotein E; PACC: preclinical Alzheimer cognitive composite.

|  | **ALFA+ cohort (N=410)** | **N** |
| --- | --- | --- |
| Age (years), Median (IQR) | 61.64 (58, 64.8) | 410 |
| Sex, n (%) |  | 410 |
| Men | 163 (39.8%) |  |
| Women | 247 (60.2%) |  |
| Education Years, Median (IQR) | 12 (11, 17) | 410 |
| CSF Aβ42/40 status*, n (%) | | 388 |
| Aβ-negative (A-) | 255 (65.7%) | |
| Aβ-positive (A+) | 133 (34.3%) | |
| APOE-ε4, n (%) | | 400 |
| non-carriers | 179 (44.8%) |  |
| carriers | 221 (55.2%) |  |
| PACC, Median (IQR) | 0.055 (-0.44, 0.46) | 410 |
| Episodic Memory, Median (IQR) | 0.063 (-0.42, 0.47) | 410 |

#### Genotyping Profiling

DNA was obtained from blood samples through a salting out protocol. Genotyping was performed with the Illumina Infinium Neuro Consortium (NeuroChip) Array (build GRCh37/hg19)^50^. The quality control procedure was performed using PLINK software v.1.9. Samples with a call rate below 98%, mismatched genetically determined sex, or excess heterozygosity (four standard deviations from the mean) were excluded. Individuals with high genetic relatedness (IBD > 0.185) were also removed. After sample-level QC, genetic variants with a minor allele frequency (MAF) below 1%, Hardy-Weinberg equilibrium p-value below 10^−6^, or missingness rates above 5% were excluded. Imputation was performed using the Michigan Imputation Server with the haplotype Reference Consortium Panel (HRC r1.1 2016) following default parameters. Further details can be found in Vilor-Tejedor *et al*^48^.

#### Plasma sample collection and proteomics assessment

Whole blood was drawn with a 20G or 21G needle gauge into a 10-ml EDTA tube (BD Hemogard, 10 ml, K2 EDTA, catalog no. 367525). Tubes were gently inverted 5–10 times and centrifuged at 2,000g for 10 min at 4 °C. The supernatant was aliquoted in volumes of 0.5 ml into sterile poly(propylene) tubes (Sarstedt Screw Cap Micro Tube, 0.5 ml, PP, ref. no. 72.730.105) and immediately frozen at −80 °C. The samples were processed at room temperature. The time between collection and freezing of plasma samples was <30 min.

Proteomic profiling of non-fasted EDTA plasma samples from 368 participants of the ALFA+ cohort was performed by SomaLogic Inc. (Boulder, CO, USA) using an aptamer-based technology (SomaScan proteomic assay). Relative protein abundances of 7,289 human protein targets were evaluated (SomaLogic v4.1 7K). Protein concentrations are quantified as relative fluorescent units (RFU). Quality control (QC) was performed by Somalogic at the sample and aptamer levels using control aptamers and calibrator samples. At the sample level, hybridization controls were used to correct for systematic variability in hybridization, and calibrators were used to correct for total signal differences caused by assay variance between individuals. The resulting hybridization scale factors and median scale factors were used to normalize data across samples within a run. Aptamer protein mapping to UniProt identifiers and gene names was provided by SomaLogic. Moreover, protein levels of significant proxies associated with cognitive performance in ALFA+ individuals were measured using immunoassay in EDTA plasma fasting samples. Human TIMP-2 Quantikine ELISA Kit #DTM200 (R&D Systems, Inc, Minneapolis, US) Elisa assay was used for plasma TIMP2 measurement, following manufacturer’s instructions, at the Clinical Neurochemistry Laboratory, Sahlgrenska University Hospital, Mölndal, Sweden. Outliers in protein expression levels (SomaLogic and ELISA) were identified and removed based on the IQR.

#### Cognitive assessment

All participants of the ALFA+ cohort underwent a cognitive test battery for the detection of early decline. A composite to assess cognitive performance was created based on the Preclinical Alzheimer Cognitive Composite (PACC)^51^. For our study, a modified PACC composite was created by averaging the Z-scores of the following variables: 1) Total Paired Recall (TPR) and 2) Total Delayed Free Recall scores of the Memory Binding Test (MBT)^52,53^, 3) the Coding subtest of the Wechsler Adult Intelligence Scale-Fourth Edition (WAIS-IV), and 4) semantic fluency. Moreover, two additional cognitive composites to assess global episodic memory (EM) and executive function (EF) were calculated by creating Z-scores for the cognitive measures from all MBT scores and from WAIS-IV subtests (Coding, Digit Span, Visual Puzzles, Similarities and Matrix Reasoning) respectively.

### 4.4. Statistical analysis

#### Proteomic-based genetic scores quantification

We obtained summary statistics data from Pietzner et al.^54^ on protein-quantitative trait *loci* (pQTLs) linked to plasma levels of the proteins identified through a systematic review. The study encompassed a cohort of 10,708 middle-aged, healthy individuals of European descent from the Fenland study, where the SomaScan (SomaLogic) platform was used to measure plasma levels of 4,775 distinct proteins.

We then computed protein genetic scores (protPRS) using PRSice version 2^55^. PRSice computes protPRS by summing all pQTL alleles carried by participants, weighting them by the pQTL allele effect size estimated in the Pietzner et al. study, and normalizing the score by the total number of pQTLs identified for each protein. The pQTLs included in the protPRS computations were those at a suggestive genome-wide level of significance (p-value<5×10^-5^). A total of 13 protPRSs were computed for 11 plasma proteins; 6 protPRSs for factors associated with brain aging and an additional 6 for factors linked to brain rejuvenation in mice. Summary statistics data of pQLTs associated with CCL11 were not available in the Pietzner. et al study, thus the protPRS of CCL11 was not computed. For the proteins haptoglobin (HP) and Brain-Derived Neurotrophic Factor (BDNF), the summary statistics was available for two different Somalogic measurements (two different aptamers were used), thus two different protPRSs for each of the protein have been computed (BDNF 1-2, HP 1-2). protPRSs were calculated in representative genetic variants per linkage disequilibrium block (LD) (clumped variants), using a cut-off for LD of r2 > 0.1 in a 250-kb window. Results were displayed at a restrictive threshold, 5×10^-5^. Additional information regarding the number of pQTLs included in each protPRSs are reported in the Supplementary Table 7.

#### Validation of genetic proxies of protein levels

To validate the utility of protPRSs as genetic proxies for plasma protein levels, we employed linear regression models in two independent cohorts (Knight-ADRC and ALFA+). In each cohort, protPRSs were used as predictors against individual plasma protein levels as outcomes, to provide a replication of the predictability of these genetic scores. All models were adjusted for age and sex to control for basic demographic variations. We further refined our analysis through stratified assessments based on sex, Aβ status, and *APOE-*ε4 carriership. Additionally, to ensure uniformity and comparability in our analysis, we standardized all protPRSs via z-scoring and we also scaled the protein levels primarily to enhance the clarity of visual representations in the outputs.

#### Associations of protein genetic scores with cognitive composites

To evaluate the associations between protPRSs and cognitive performance, we conducted linear regression analyses across cognitive composites. Each protPRS served as a predictor with cognitive composites as outcomes. Our models were adjusted for age, sex, and years of education to account for potential confounding effects. To enable direct comparisons of beta estimates, protPRSs were standardized using z-scoring techniques.

Further, we conducted stratified analyses to investigate the potential influence of sex, Aβ status (defined as CSF Aβ42/40 ratios below 0.071)^49^, and *APOE-*ε4 carriership on the association between protPRSs and cognitive outcomes. The statistical significance of our findings was evaluated at various levels: nominal significance, adjustment for differences between two experimental aging groups (aging/rejuvenating), and corrections for multiple testing using the False Discovery Rate (FDR) method. Results were reported with corresponding significance thresholds (*p<0.05, **p<0.025, ***FDR-corrected p-value <0.05). For the significant genetic proxies, we validated the aptamer recognition of the respective protein by non-parametric Spearman correlation test between Somalogic protein levels and those measured by ELISA immunoassay.

#### TIMP2 protPRS annotation and gene set enrichment analysis

Significant plasma pQTLs (P<5×10^-5^), included in the TIMP2-protPRS after clumping, were annotated using the Ensembl Variant Effect Predictor release 111^56^, based on GENCODE v19 transcripts for the GRCh37 human genome assembly^57^ (Supplementary Table 9-10). To further obtain insight into putative biological mechanisms, gene set enrichment analysis of annotated genes was conducted using *clusterProfiler*^58^ (FDR-adjusted p-value ≤ 0.05). Furthermore, GTEx v8 eQTL data was used to establish tissue-specific gene expression patterns and their relevance to the TIMP2 protein levels measured in our pQTL analysis (GENE2FUNC function^59^ FUMA v1.5.2). By mapping eQTLs from GTEx to the genomic locations identified in our TIMP2 pQTL analysis, it was assessed whether variations that affect TIMP2 plasma protein levels also influence significant differential gene expression in a tissue-specific manner across 54 tissue types.

## Data Availability

All data produced in the present study are available upon reasonable request to the authors

## Acknowledgements

The authors express their most sincere gratitude to the participants and relatives of the ALFA and Knight ADRC studies, without whom this research would have not been possible. This publication is part of the ALFA study. Collaborators of the ALFA study are: Müge Akinci, Annabella Beteta, Raffaele Cacciaglia, Lidia Canals, Alba Cañas, Carme Deulofeu, Maria Emilio, Irene Cumplido-Mayoral, Ruth Dominguez, Karine Fauria, Sherezade Fuentes, Laura Hernández, Gema Huesa, Jordi Huguet, Laura Iglesias, Esther Jiménez, Helena Blasco, Javier Torres, David López-Martos, Paula Marne, Tania Menchón, Paula Ortiz-Romero, Marina de Diego, José Contador-Muñana, Armand González-Escalante, Anna Brugulat-Serrat, Eleni Palpatzis, Wiesje Pelkmans, Albina Polo, Sandra Pradas, Mahnaz Shekari, Lluís Solsona, Anna Soteras, Núria Tort-Colet, and Marc Vilanova. Funding and support for the plasma proteomics analysis was provided by Gates Ventures. We would like to thank Kaj Blennow and Henrik Zetterberg for their support on measuring TIMP2 levels. We thank Roche Diagnostics International for providing the kits to measure CSF biomarkers of ALFA+ participants.

## Consent Statement

The ALFA study was conducted in accordance with the directives of the Spanish Law 14/2007, of July 3, on Biomedical Research (Ley 14/2007 de Investigación Biomédica). The ALFA study protocol was approved by the Independent Ethics Committee Parc de Salut Mar Barcelona and registered at Clinicaltrials.gov (Identifier: NCT02485730). All participants accepted the study procedures by signing the study’s informed consent form that had also been approved by the same Institutional Review Board. The ethics committee and institutional review board of Washington University in St. Louis School of Medicine approved this study. All participants provided informed consent for all data used in the present study.

## Supplementary Materials

## 1. Supplementary Figures

S Figure 1. Systematic review workflow. Systematic review process, guiding the selection and evaluation of relevant studies to identify aging and rejuvenating factors in mice.

S Figure 2. Plots of protPRSs association with plasma protein levels in the Knight-ADRC cohort. The figure reports the plots of the association model of protPRSs with plasma protein levels in the Knight-ADRC cohort. Significant associations are indicated by an asterisk next to the protein name.

S Figure 3. Plots of protPRSs association with plasma protein levels in the ALFA+ cohort. The figure reports the plots of the association model of protPRSs with plasma protein levels in the ALFA+ cohort. Significant associations are indicated by an asterisk next to the protein name.

S Figure 4. Correlation between SomaScan and ELISA immunoassay TIMP2 plasma levels in ALFA+. The plot represents the correlation (Spearman correlation test) between TIMP2 levels assessed via ELISA immunoassay and those derived from SomaScan technology.

S Figure 5. Correlation between TIMP2 protPRS and ELISA immunoassay TIMP2 plasma levels in ALFA+. The plot represents the correlation (Spearman correlation test) between TIMP2 levels assessed via ELISA immunoassay and the computed TIMP2 protPRS.

S Figure 6. Mapping of pQTLs for TIMP2 plasma levels. Functional annotation of genetic variants associated with TIMP2 plasma pQTLs, conducted using FUMA. Only pQTLs that remained significant after Bonferroni correction were considered. The functional consequences of SNPs were evaluated using Ensembl genes (build 85). Asterisks denote significant differences in proportions between TIMP2 plasma pQTLs and the reference panel (*P < 0.05).

S Figure 7. Enrichment analysis of the annotated SNPs included in the TIMP2 protPRS after clumping.

S Figure 8. Tissue-dependent regulation of genes associated with TIMP2 pQTLs. Red bars indicate tissues with statistically significant regulation.

## 2. Supplementary Tables

S Table 1. Systematic review results.

S Table 2. Papers included in the systematic review

S Table 3. pQTLs summary statistic data from Pietzner et al.

S Table 4. protPRSs association with PACC in the ALFA+ cohort

S Table 5. protPRSs association with EM in the ALFA+ cohort

S Table 6. Plasma TIMP2 protein (ELISA and SomaScan) association with PACC and EM

S Table 7. pQTLs included in the protPRSs

S Table 8. SNPs included in the TIMP2 protPRS of ALFA+ after clumping

S Table 9. TIMP2 protPRS annotated gene list

