## Supplementary figures and images for "Polygenic proxies of age-related plasma protein levels reveal TIMP2 role in cognitive performance"

### S_Figure_1.png

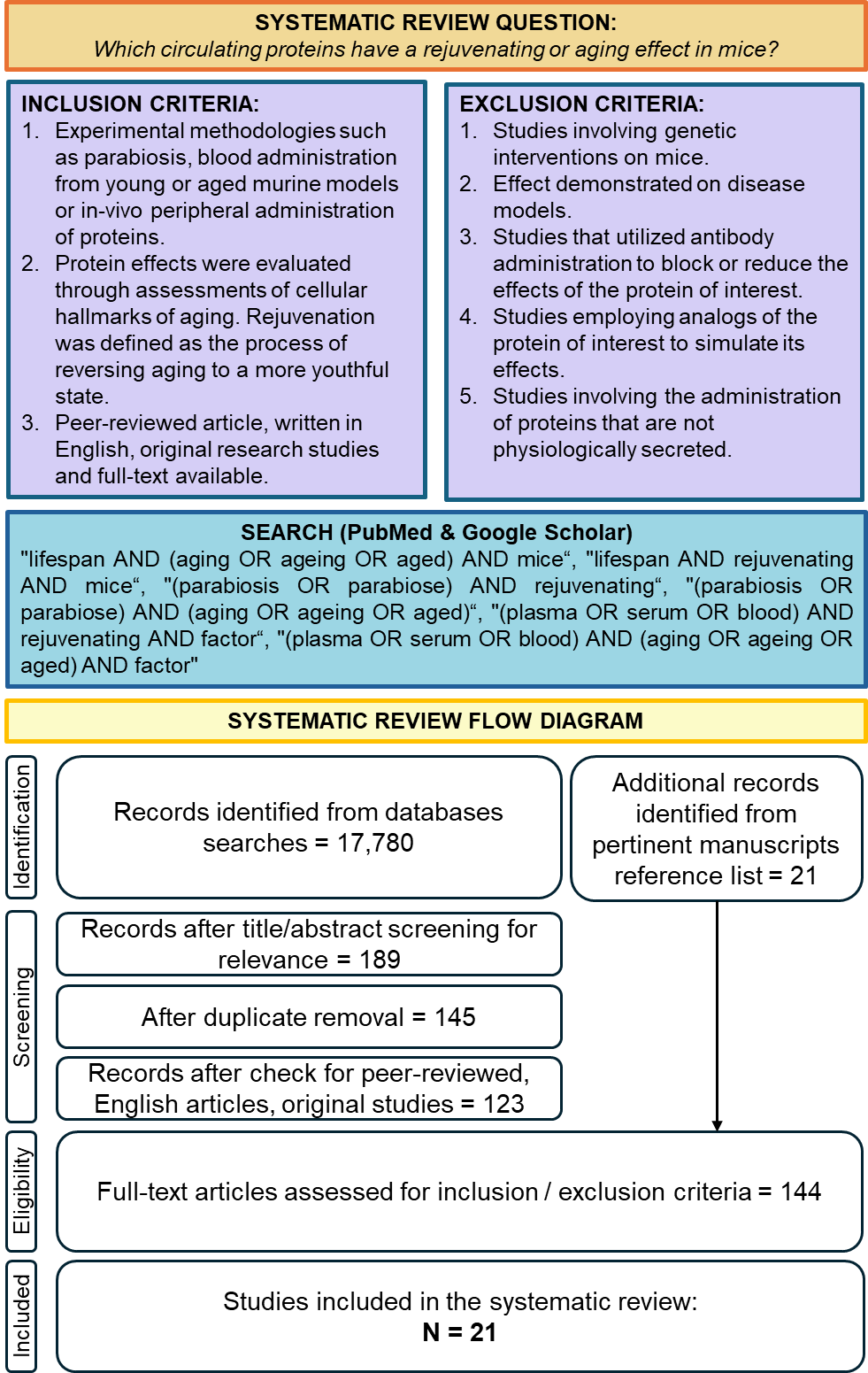

### S_Figure_2.png

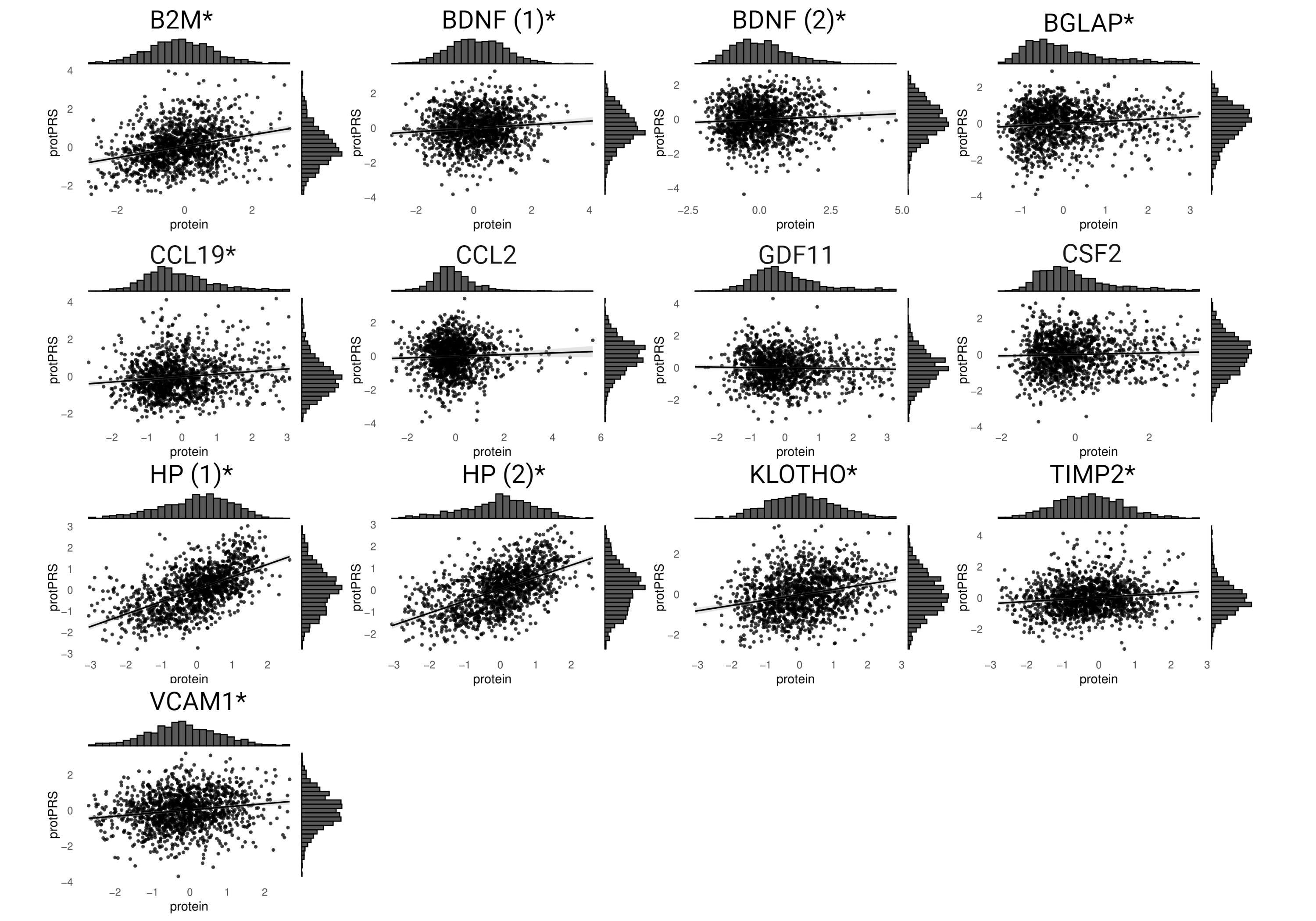

### S_Figure_3.png

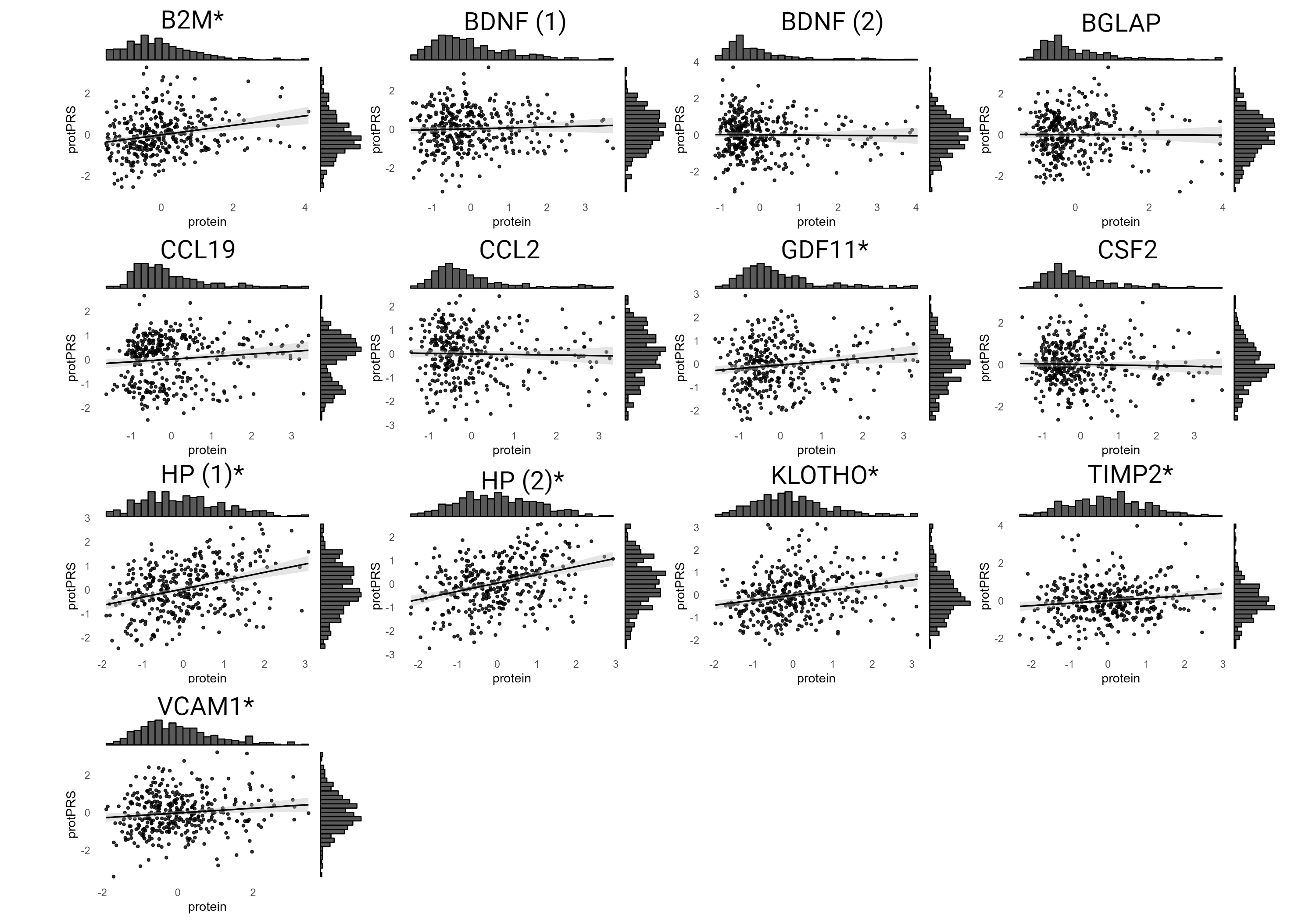

### S_Figure_4.png

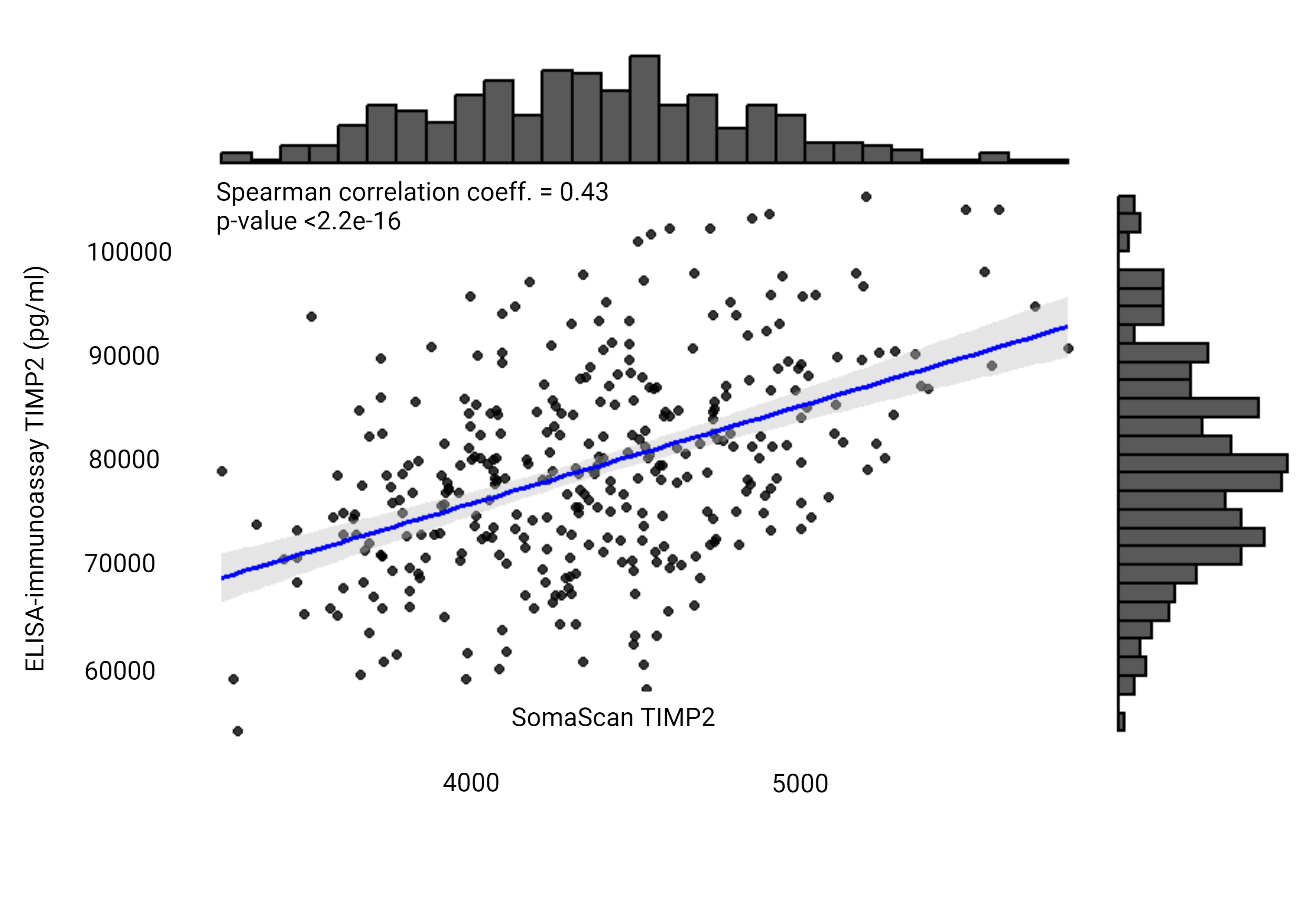

### S_Figure_5.png

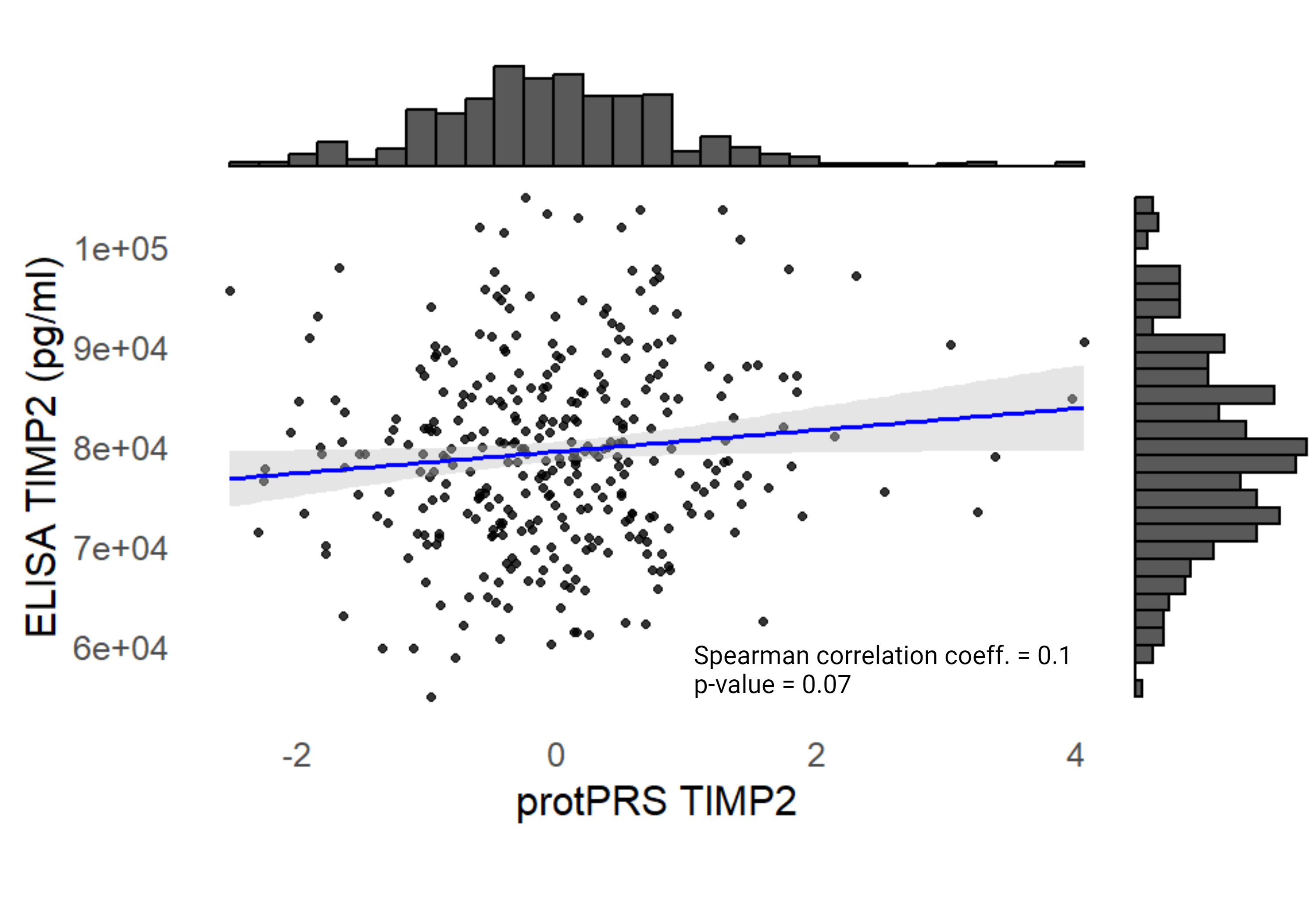

### S_Figure_6_rev.png

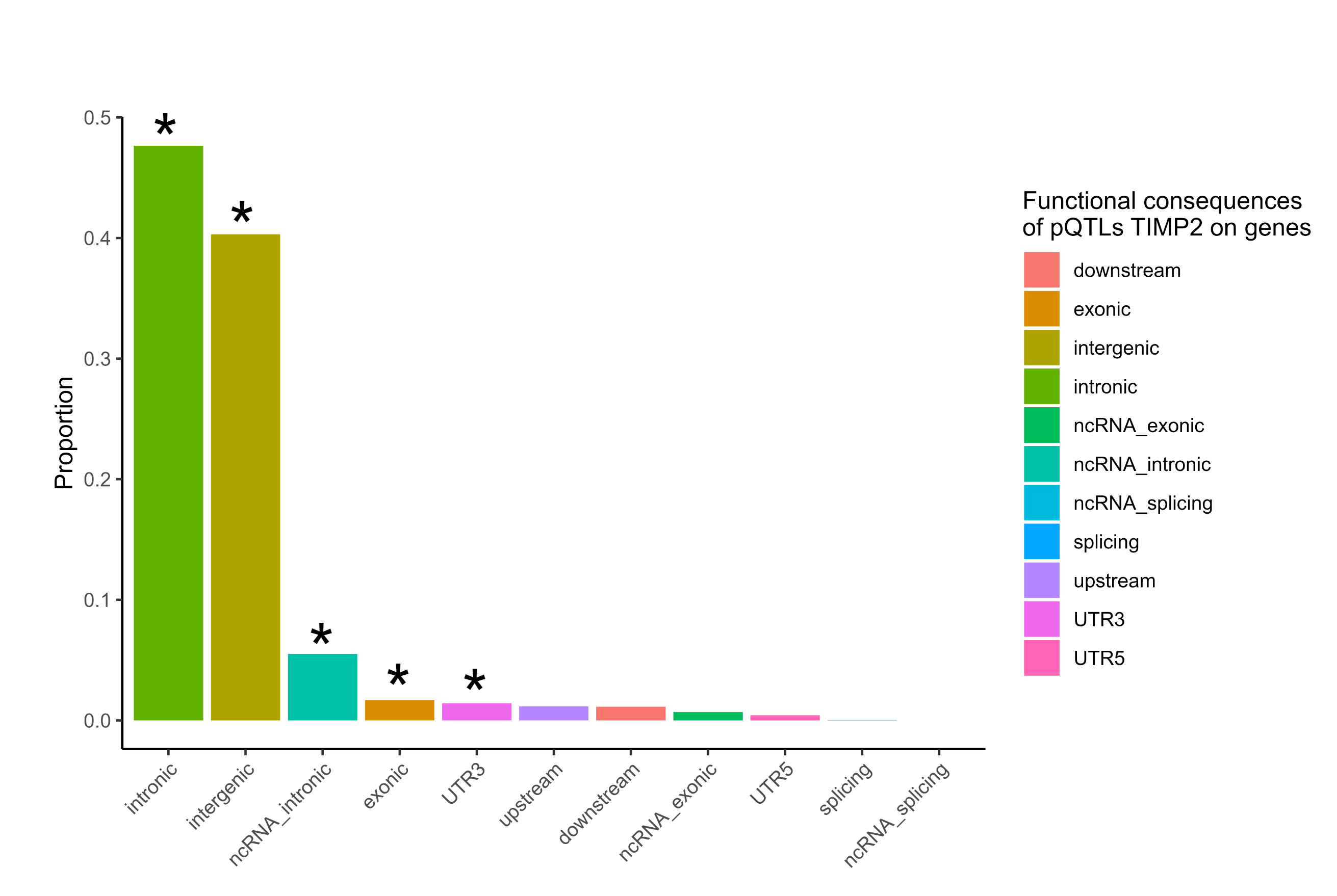

### S_Figure_7.png

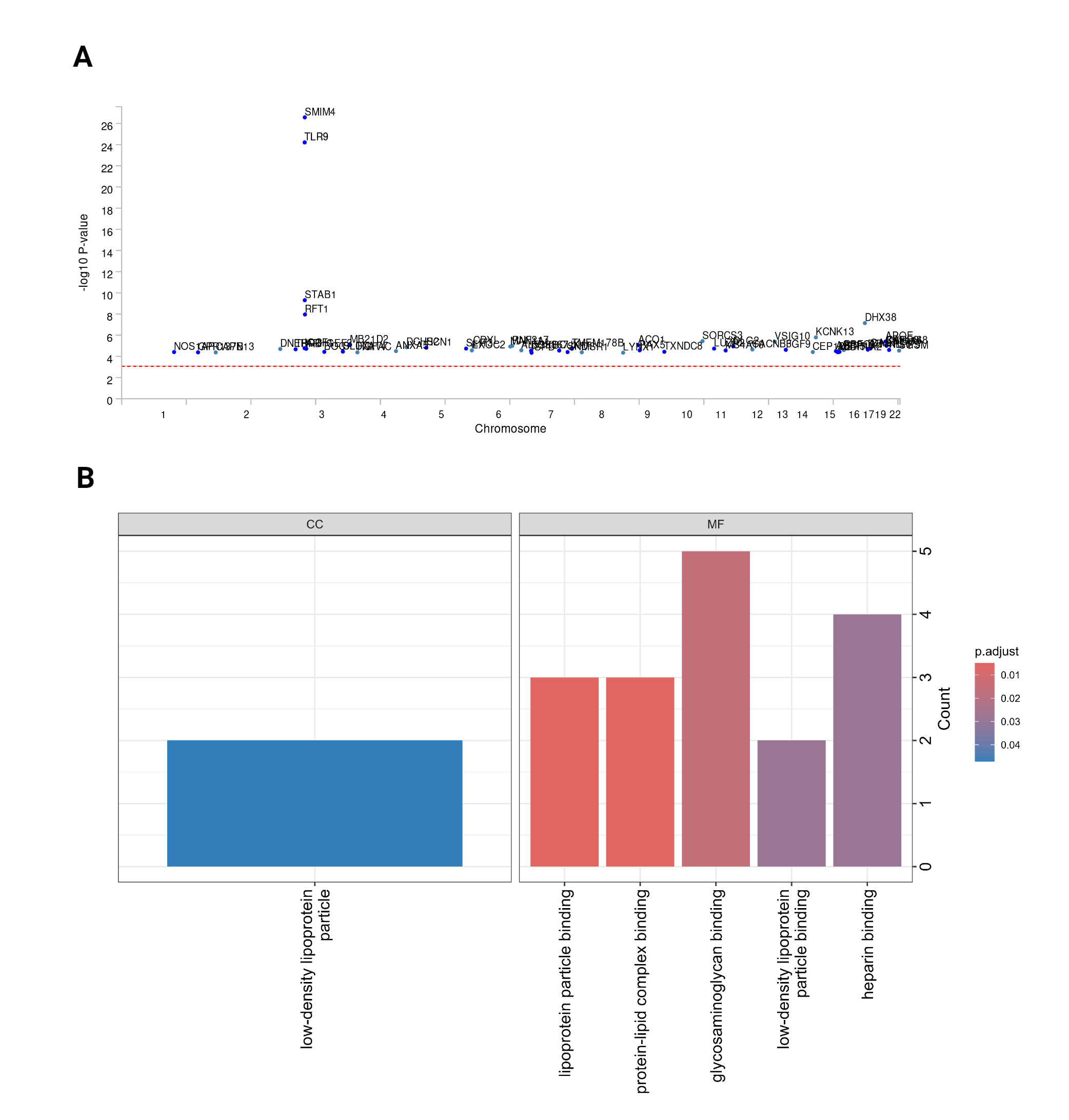

### S_Figure_8.png

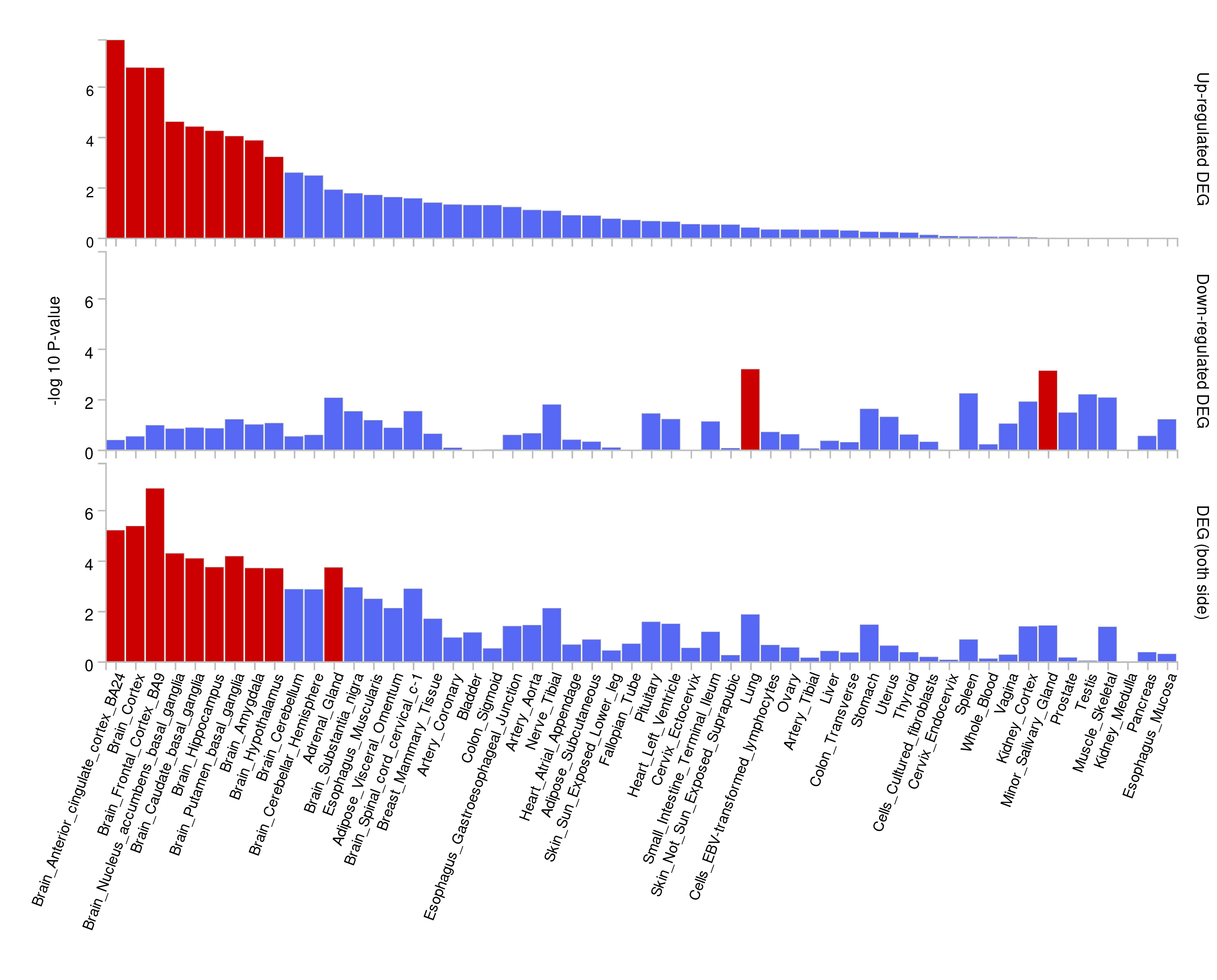
